# Evaluation of standard and enhanced quality improvement methods to increase the uptake of magnesium sulfate in preterm deliveries for the prevention of neurodisability (PReCePT Study): a cluster randomized controlled trial

**DOI:** 10.1101/2022.05.20.22275244

**Authors:** Hannah B Edwards, Maria Theresa Redaniel, Carlos Sillero-Rejon, Christalla Pithara-McKeown, Ruta Margelyte, Tracey Stone, Tim J Peters, William Hollingworth, Hugh McLeod, Pippa Craggs, Elizabeth M Hill, Sabi Redwood, Emma Treloar, Jenny L Donovan, Brent C Opmeer, Karen Luyt

**Affiliations:** National Institute for Health and Care Research Applied Research Collaboration West (NIHR ARC West) at University Hospitals Bristol and Weston NHS Foundation Trust. Whitefriars Level 9, Lewins Mead, Bristol, BS1 2NT, UK; Population Health Sciences, Bristol Medical School, University of Bristol. 5 Tyndall Avenue, Bristol, BS8 1UD, UK; Research and Innovation, University Hospitals Bristol and Weston NHS Foundation Trust. Level 3, Upper Maudlin Street, Bristol, BS2 8AE, Bristol, UK; St. Michael’s Hospital, University Hospitals Bristol and Weston NHS Foundation Trust. Southwell Street, Bristol, BS2 8EG, UK; West of England Academic Health Science Network. 360 Bristol – Three Six Zero, Marlborough Street, Bristol, BS1 3NX, UK; University of Oxford, Oxford University Hospitals NHS Foundation Trust, Oxford Academic Health Science Network. Whitehead Building, 1 Robert Robinson Avenue, Oxford Science Park, Oxford, OX4 4GA, UK; Translational Health Sciences, Bristol Medical School, University of Bristol. 5 Tyndall Avenue, Bristol, BS8 1UD, UK

**Author notes:** **Corresponding author:** Hannah Edwards, National Institute for Health and Care Research Applied Research Collaboration West (NIHR ARC West), 9th Floor, Whitefriars, Lewins Mead, Bristol, BS1 2NT, UK. contributed equally to the paper (Hannah B Edwards, Maria Theresa Redaniel, Carlos Sillero-Rejon, Christalla Pithara-McKeown, Ruta Margelyte).

**Keywords:** Obstetrics, Midwifery, Pregnancy, Pregnant Women, Infant, Premature, Infant, Extremely Premature, Cerebral Palsy, Magnesium Sulfate, National Health Programs, Quality Improvement, Quality of Health Care, Randomized Controlled Trials, Cluster Analysis

## Abstract

**Objective:** To compare the impact of the National PReCePT Programme (NPP) versus an enhanced Quality Improvement (QI) support programme in improving magnesium sulfate (MgSO_4_) uptake in English maternity units.

**Design:** Unblinded cluster randomised controlled trial.

**Setting:** England, Academic Health Sciences Network (AHSN), 2018.

**Participants:** Maternity units with ≥10 preterm deliveries annually and MgSO_4_ uptake ≤70%. 40 maternity units (27 NPP, 13 enhanced support) were included (randomisation stratified by MgSO_4_ uptake).

**Interventions:** NHS England commissioned the NPP to increase MgSO_4_ uptake in very preterm deliveries to reduce risk of cerebral palsy. NPP maternity units received PReCePT QI materials, regional support, and midwife backfill funding. Enhanced support units received this plus extra backfill funding and unit-level QI coaching.

**Outcome measures:** MgSO_4_ uptake post-implementation was compared between groups using routine data and multivariable linear regression. Net monetary benefit was estimated, based on implementation costs, lifetime quality-adjusted life-years and societal costs. The implementation process was assessed through qualitative process evaluation.

**Results:** MgSO_4_ uptake increased in all units, with no evidence of difference between groups (0.84 percentage points lower uptake in the enhanced group, 95% Confidence Interval -5.03 to 3.35 percentage points). The probability of enhanced support being cost-effective was <30%. NPP midwives allocated more than their funded hours. Units varied in support required to successfully implement the intervention. Enhanced support units reported better understanding, engagement, and perinatal teamwork.

**Conclusion:** PReCePT improved MgSO_4_ uptake in all maternity units. Enhanced support did not further improve uptake but may improve teamwork, and more accurately represented the time needed for implementation. Targeted enhanced support, sustainability of improvements and the possible indirect benefits of stronger teamwork associated with enhanced support should be explored further.

**Trial registration:** ISRCTN 40938673 (https://www.isrctn.com/ISRCTN40938673)

**WHAT IS ALREADY KNOWN ON THIS TOPIC:**

- Despite long-standing evidence that Magnesium Sulfate (MgSO_4_) confers fetal neuroprotection and reduces risk of cerebral palsy in very preterm babies, by 2017 only two-thirds of eligible women in England were receiving it, with wide regional variation.
- The pilot PReCePT (Prevention of Cerebral Palsy in preterm labour) Quality Improvement (QI) study appeared to effectively accelerate uptake of MgSO_4,_ and a version of this support model was rolled-out nationwide in 2018.

*WHAT THIS STUDY ADDS:*

- PReCePT improved MgSO_4_ uptake in all maternity units, and the full (‘enhanced’) support model did not appear to improve uptake beyond the achievements of the standard support model used in the National PReCePT Programme. However, enhanced support may be associated with improved perinatal team working, and the funding more accurately represented the staff time needed for implementation.

*HOW THIS STUDY MIGHT AFFECT RESEARCH, PRACTICE OR POLICY:*

- PReCePT may serve as a blueprint for other improvement programs to accelerate uptake of evidence-based interventions, and future studies should consider the potential for indirect but far-reaching benefits to staff and patients.

## INTRODUCTION

Neurodisabilities due to preterm birth, including cerebral palsy (CP), represents a significant burden for individuals, families,(1) and healthcare services.(2-4) Antenatal magnesium sulfate (MgSO_4_) reduces the risk of CP in preterm births by around 30%.(5) A dose costs approximately £1(6) with estimated lifetime societal savings of approximately £1M per case of CP avoided.(7)

Since 2015, the UK National Institute for Health and Care Excellence (NICE) has recommended administration of MgSO_4_ in preterm deliveries(8) and non-compliance is considered sub-optimal care. Yet by 2017, only 64% of eligible women (<30 weeks gestation) were receiving MgSO_4._(9)

The PReCePT (Preventing Cerebral Palsy in Pre-Term labour) Quality Improvement (QI) intervention was developed to improve maternity staff awareness and increase MgSO_4_ uptake. The pilot study (five maternity units) improved MgSO_4_ uptake from 21% in 2012-2013 to 88% in 2015.(10) The National PReCePT Programme (NPP) scaled-up the intervention and it was rolled-out across English maternity units, led by regional Academic Health Science Networks (AHSNs) to increase MgSO_4_ uptake to 85% by 2020(11).

A cluster randomised controlled trial (cRCT) was nested within the NPP. It evaluated the effectiveness and cost-effectiveness of the enhanced support model compared to the standard NPP support model(12). A qualitative process evaluation was conducted to understand the implementation process in both groups.(13)

## METHODS

### Trial Design

This unblinded nested cRCT was set in NHS England maternity units. NPP (control) units received standard NPP support including PReCePT QI guide and toolkit resources (e.g., preterm labour proforma, staff training presentations, parent leaflet, posters for the unit, learning log), regional AHSN-level support, and up to 90 hours funded backfill for a midwife ‘champion’ to lead implementation. Enhanced support (intervention) units received this plus unit-level QI coaching for the lead midwife, obstetrician and neonatologist, additional 90 hours midwife backfill funding, approximately 104 hours backfill funding for the local obstetrician/neonatologist lead, team access to learning and celebration events, and a computer tablet for micro-coaching staff (Supplementary file 1: Description of trial groups). The trial was embedded within the NPP and aligned with its timeframe of two waves (Figure 1). After randomisation, implementation ran for nine months with a further nine months’ follow-up.

**Figure 1.**
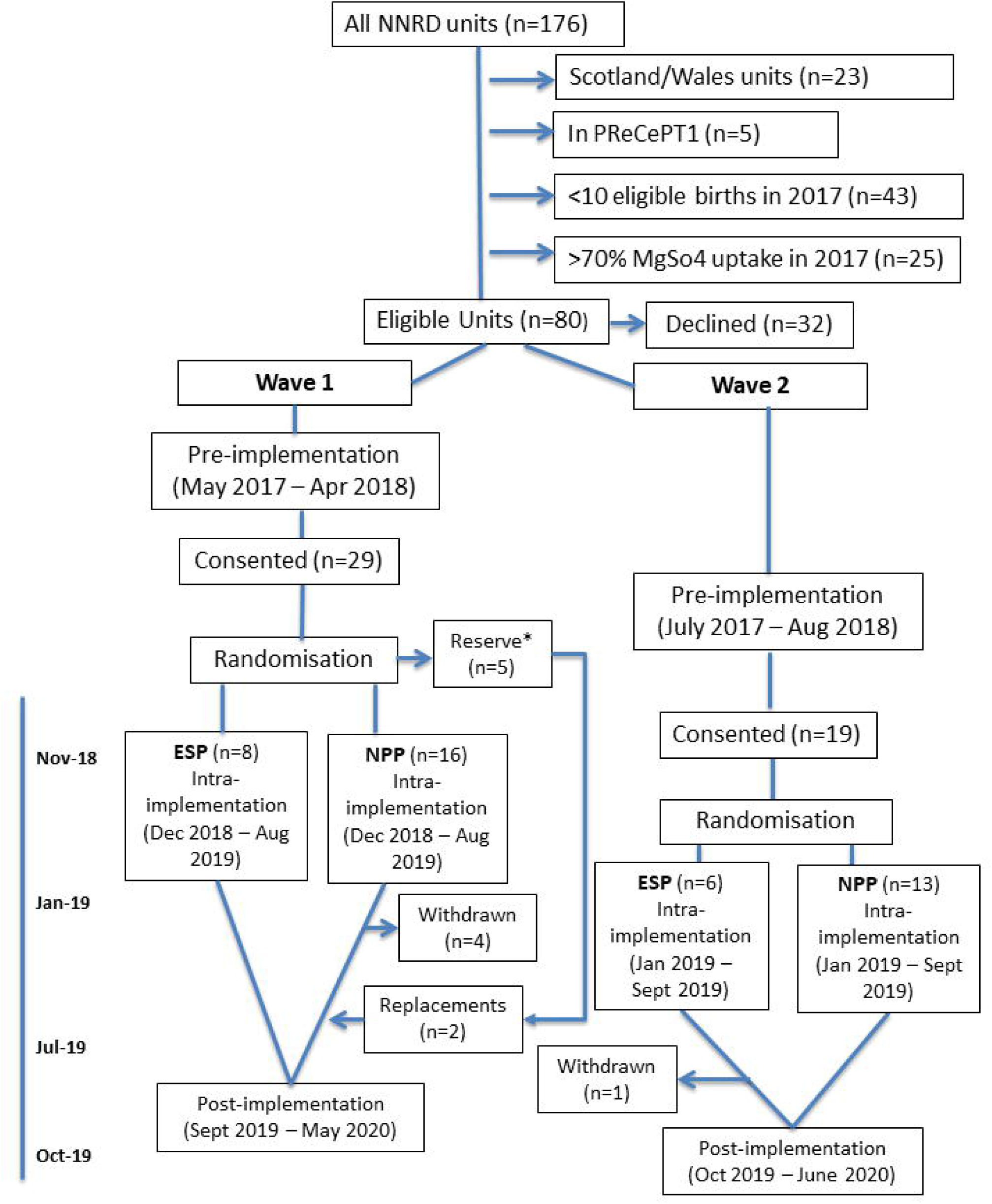

### Eligibility criteria

Maternity units in England participating in the NPP with ≥10 preterm (<30 weeks gestation) deliveries annually and MgSO_4_ uptake ≤70% were eligible. Eligibility was assessed from 2017 UK National Neonatal Research Database (NNRD) data, in units that expressed an interest in participating. PReCePT pilot study units were excluded.

### Outcomes

The primary outcome was the unit-level proportion of eligible women receiving MgSO_4_ post-implementation. Unit-level assessment of outcome was appropriate given that the intervention was delivered at the unit-level, and clustering effects were likely. Secondary outcomes included uptake and MgSO_4_ data completeness over time, reasons MgSO_4_ was not given, and costs and cost-effectiveness (incremental net monetary benefit) from a societal perspective over the lifetime of a preterm baby. A qualitative process evaluation assessed the implementation process, fidelity, and local adaptations to elucidate similarities and differences between study arms, and any unintended outcomes.

### Sample size and randomisation

At the time of study design, the background population was 153 English maternity units. There was limited data for power calculation parameter assumptions, but at the design stage, data from the 2016 National Neonatal Audit Programme (NNAP) data, the pilot study, and clinical assumptions indicated an anticipated baseline MgSO_4_ uptake in the control arm of 38%, uptake in the enhanced support arm of 80%, and a high intraclass correlation coefficient (ICC) of up to 0.67 (formulae for cluster trials taken from the literature(14)). To detect a difference of 40 percentage points in uptake between groups, that would not only be clearly important but also feasible as suggested by the available data, with a 2-sided 5% significance level, 80% power, ICC=0.67, coefficient of variation for cluster size=0.48, and a 1:2 randomisation ratio, 11 intervention and 22 control units were needed.

Units were stratified by 2017 MgSO_4_ uptake rates (0-39.9%, 40-49.9%, 50-59.9%, and 60-70.9% uptake). Taking into account these four strata and two implementation waves, the target sample size was increased to 48.

Randomisation was performed in two tranches, in line with the two waves of implementation. It was performed with Stata command *stratarand* and carried out by a statistician independent of the trial and NPP (Supplementary file 2: Randomisation).

The nature of the interventions made it impossible to conceal allocation from maternity staff. The unequal randomization ratio also made it difficult to conceal allocation from research staff performing the analysis.

### Data collection

We used pseudonymized patient-level data from the NNRD.(15, 16) Baseline data were collected for the 12 months pre-implementation. Index of Multiple Deprivation (IMD) data was derived from published data for each Lower Super Output Area.(17) Data on number of beds and staff, amount of time spent on PReCePT-related activities, and previous QI experience were collected via questionnaires completed by unit lead midwives. Cost data was supplied by the NPP team.

For the process evaluation, criterion-based sampling (trial arm, annual number of births, baseline MgSO_4_ uptake, recent Care Quality Commission (CQC) ratings on leadership and patient safety) was used to select units for qualitative interviews. Implementers (unit lead midwife, obstetrician and neonatologist) were invited to participate in a semi-structured telephone interview near the end of the implementation period. Interviews explored: experiences of QI activities, staff engagement, perceived leadership support, and contextual factors (professional/ cultural issues, organisational changes, staff shortages, impact of Coronavirus-19) (Supplementary file 4: Qualitative interviews topic guide). Written informed unit and individual consent were obtained and interviews were audio-recorded and transcribed.

### Data analyses

#### Primary outcome

MgSO_4_ uptake was defined as the number of mothers given MgSO_4_ divided by the total number of eligible mothers, excluding missing values from the denominator, expressed as a percentage.(9) Baby-level demographic descriptions included all babies. In all other analyses, we only included data for singletons and the first-born of multiples (for consistency with nationally reported audit data). Where only one baby had a record for MgSO_4_, the missing MgSO_4_ status of the other multiples was recoded to match that for their twin/triplet with a record. For babies with conflicting records, we recorded MgSO_4_ as given.

Linear regression was used to assess differences in MgSO_4_ uptake between trial arms post-implementation, adjusted for pre-implementation uptake. The model was weighted on the number of births in each unit and used robust standard errors. Sensitivity analyses adjusted for factors by which the trial arms differed appreciably pre-implementation.

#### Secondary outcomes

Controlled interrupted time series (ITS) analysis using segmented linear regression was used to model differences in trends in uptake and missing MgSO_4_ data, over three time periods: pre, intra- and post-implementation. Newey-West standard errors (with one lag) were estimated by ordinary least-squares regression and used to handle autocorrelation in the model. Differences in slope (indicating trend) and intercept (value of MgSO_4_ uptake) between trial arms, as well as differences across the time periods, were described in the model.

#### Cost-effectiveness

Implementation costs in both arms included management, AHSN support, and midwife backfill. Additional enhanced support costs were incurred by extra clinical and midwife backfill, unit-level QI coaching, and learning events. Staff time was costed using national salary data. Mean implementation cost per baby was calculated as the mean implementation cost per unit divided by the total number of babies eligible for MgSO_4_ per unit delivered during the implementation and follow-up period. Mean staff time per week spent on MgSO_4_ activities was estimated from questionnaires completed for the month before starting the QI and each intra-implementation month.

Decision tree analysis estimated enhanced support net monetary benefit using a lifetime horizon and societal perspective. Model parameters were based on trial data for implementation costs and MgSO_4_ uptake, literature estimates(7) for lifetime gains in quality-adjusted life-years (QALYs), and societal cost savings from MgSO_4_ treatment for imminent and threatened preterm births (Supplementary file 3, Table S3). Babies delivered by caesarean section were defined as imminent births (certain to occur within 24 hours) and all others as threatened. We used a £20,000 per QALY gained willingness-to-pay threshold.(18)

The probability of MgSO_4_ treatment in the enhanced support group compared with NPP was estimated using a multilevel logistic regression model clustered at unit-level to determine the odds ratio of imminent and threatened babies having received MgSO_4_ during the 18-month implementation and follow-up period, adjusted for baseline uptake. For this analysis only, babies with missing MgSO_4_ treatment records were assumed to have not received treatment We conducted a probabilistic analysis using Monte Carlo simulation with 10,000 samples drawn from the parameter distributions. Incremental costs and effects were plotted on the cost-effectiveness plane and a cost-effectiveness acceptability curve was plotted for willingness-to-pay thresholds from £0 to £100,000 per QALY gained. This analysis accounts for parameter uncertainties by drawing samples at random from parameter-specific probability distributions. (Supplementary file 3, Figure S1)

#### Process evaluation

Semi-structured interviews were analysed using the framework method.(19) The matrix output, using rows, columns and ‘cells’ of summarised data, facilitated analysis by case (for example, site, professional group, or individual) and by code (summarised data in relation to a particular theme such as intervention fidelity). This allowed comparison of data across and within cases to inform understanding of the implementation processes by which this complex intervention is operationalised, embedded, and sustained in practice. Analysis focused on aspects of individual and collective behaviour shown to be important in implementation processes.(20)

## RESULTS

Applying eligibility criteria to all 153 English maternity units left 80 units (52% of the total population) as potentially eligible participants. Of these, 48 (60%) were randomised. Due to changes in some units’ readiness to start, and the need to balance randomisation between tranches and strata, 40 units were included (13 enhanced support, 27 standard NPP, Figure 1). (See also Supplementary file 2: Randomisation; Supplementary Table S1). This covered 2,962 babies born to 2,597 mothers in the pre- and post-implementation periods. Trial arms were comparable at baseline. Enhanced support units saw more white British mothers and more mothers from socio-economically deprived areas. Standard support units had more experience with QI (Table 1).

**Table 1.** Socio-demographic and clinical characteristics of mothers and babies by trial arm.

| Characteristic | Enhanced support<br>(n=13 units) |  | NPP support<br>(n=27 units) |  |
| --- | --- | --- | --- | --- |
|  | Pre-implementation | Post-implementation | Pre-implementation | Post-implementation |
| **Babies** |  |  |  |  |
| Number of babies | 596 | 374 | 1148 | 844 |
| Male sex (N, %) | 333 (55.9) | 207 (55.4) | 624 (54.4) | 465 (55.6) |
| Median gestational age (weeks, median, IQR) | 28.3 (26.6 – 29.9) | 28.6 (26.4 - 30.0) | 28.6 (26.6 – 30.0) | 28.6 (26.6 – 30.0) |
| Median birthweight (g, median, IQR) | 1057.5 (800.5 – 1300) | 1089.5 (806 – 1365) | 1065 (825 – 1335) | 1057.5 (840 – 1330) |
| Number born in multiples (N, %) | 136 (22.8) | 97 (25.9) | 292 (25.4) | 194 (23.0) |
| **Mothers** |  |  |  |  |
| Number of mothers | 530 | 328 | 997 | 742 |
| Median maternal age (years, median, IQR) | 30 (25 – 34) | 30 (26 – 35) | 31 (26 – 35) | 31 (26 – 36) |
| White ethnicity (N, %) | 312 (72.2) | 167 (68.4) | 452 (56.8) | 333 (58.3) |
| IMD quintile (N, %) |  |  |  |  |
| 1 – most deprived | 199 (38.8) | 135 (42.2) | 306 (31.1) | 242 (33.4) |
| 2 | 114 (22.2) | 71 (22.2) | 252 (25.6) | 152 (21.0) |
| 3 | 73 (14.2) | 50 (15.6) | 189 (19.2) | 156 (21.6) |
| 4 | 66 (12.9) | 35 (10.9) | 140 (14.2) | 105 (14.5) |
| 5 – least deprived | 61 (11.9) | 29 (9.1) | 97 (9.9) | 69 (9.5) |
| Caesarean delivery (N, %) | 287 (61.1) | 176 (55.2) | 581 (60.0) | 429 (60.0) |
| Had pregnancy-induced hypertension (N, %) | 26 (5.0) | 19 (5.9) | 52 (5.2) | 36 (4.9) |
| Antenatal steroids given (N, %) | 479 (91.4) | 303 (93.2) | 919 (92.2) | 691 (93.6) |
| **Maternity units** |  |  |  |  |
| Level of birth unit (N mothers, %) |  |  |  |  |
| Special Care Unit (SCU) / High Dependency Unit (HDU) | 191 (36.4) | 110 (33.8) | 366 (36.7) | 265 (35.9) |
| Neonatal Intensive Care Unit (NICU) | 334 (63.6) | 215 (66.2) | 632 (63.3) | 473 (64.1) |
| Number of staff per unit (median, IQR) |  |  |  |  |
| Midwives (bands 5-8c) | 83 (60 – 166) | Only collected pre-implementation | 81 (57 – 161) | Only collected pre-implementation |
| Consultants | 15 (11 – 22) |  | 14 (9 – 24) |  |
| Delivery suite beds per unit (median, IQR) | 10 (8 – 12) |  | 12 (9 – 15) |  |
| Have previous QI experience (N, %) | 6 (46.15) |  | 19 (70.37) |  |

### Primary outcome

Mean MgSO_4_ uptake in the 12 months pre-implementation was 68.1% in NPP units, and 64.3% in enhanced support units. This increased to 83.7% and 84.8% respectively in the 12 months post-implementation (Table 2). After adjusting for pre-implementation uptake, there was no evidence of a difference in uptake between trial arms (0.84 percentage points lower uptake in the enhanced support versus NPP arms, 95% CI -5.03 to 3.35 percentage points, p=0.687).

**Table 2.** MgSO_4_ uptake in maternity units, by trial arm and study periods.

| Variable | Enhanced support |  | Standard NPPsupport |  |
| --- | --- | --- | --- | --- |
|  | Pre-implementation | Post-implementation | Pre-implementation | Post-implementation |
| **Crude proportion uptake and missing:** |  |  |  |  |
| Total number of eligible births <sup>0</sup> | 525 | 325 | 998 | 738 |
| Mothers **given** MgSO<sub>4</sub> (N, %) | 357 (68.0%) | 270 (83.1%) | 675 (67.6%) | 607 (82.2%) |
| Mothers **not given** MgSO<sub>4</sub> (N, %) | 143 (27.2%) | 51 (15.7%) | 279 (28.0%) | 109 (14.8%) |
| With MgSO<sub>4</sub> data missing (N, %) | 25 (4.8%) | 4 (1.2%) | 44 (4.4%) | 22 (3.0%) |
| **Overall proportion uptake:**<sup>1</sup> | 64.2% | 84.8% | 68.1% | 83.6% |

<sup>0</sup> Records on singleton births and the first-born of multiples included in the analysis
<sup>1</sup> Total uptake over the follow up and baseline periods. Uptake proportion excluding missing from denominator and only including singletons and first-born of multiples. Calculated from unit-level proportions per time period

Sensitivity analyses adjusting for factors imbalanced pre-implementation (maternal ethnicity, socio-economic deprivation, and previous QI experience) gave similar results (0.47 percentage points higher uptake in the enhanced support group, 95% CI -4.18 to 5.12 percentage points, p=0.840).

### Secondary outcomes

Trends in MgSO_4_ uptake were similar between groups. The proportion missing data for the enhanced support group decreased in the pre-implementation period compared to NPP units and slightly increased post-implementation, but these trends represent very small differences (Supplementary Figure S3). Overall, the amount of missing MgSO_4_ data reduced over the study period. The ICC was 0.019, indicating lower than expected clustering at the maternity-unit level.

### Costs and cost-effectiveness analyses

The incremental funded implementation cost was £16,869 per enhanced support unit, and £276 per preterm baby delivered (Supplementary tables S4, S6). The incremental impact of enhanced support on MgSO_4_ uptake over the 18 months implementation and follow-up was - 0.79 percentage points (95% CI -6.00 to 4.41).

From a societal lifetime perspective, probabilistic analysis showed a decrease of -0.001 QALYs (95% CI -0.009 to 0.006) and a cost increase of £315 per preterm baby delivered associated with the enhanced support model. This generated a net monetary loss of £340 for a willingness to pay threshold of £20,000, indicating that enhanced support was not cost-effective compared to the standard NPP model (Table 3). The probability of enhanced support being cost-effective was less than 30% across the range of plausible willingness-to-pay thresholds. (Supplementary Figure S2).

**Table 3.**
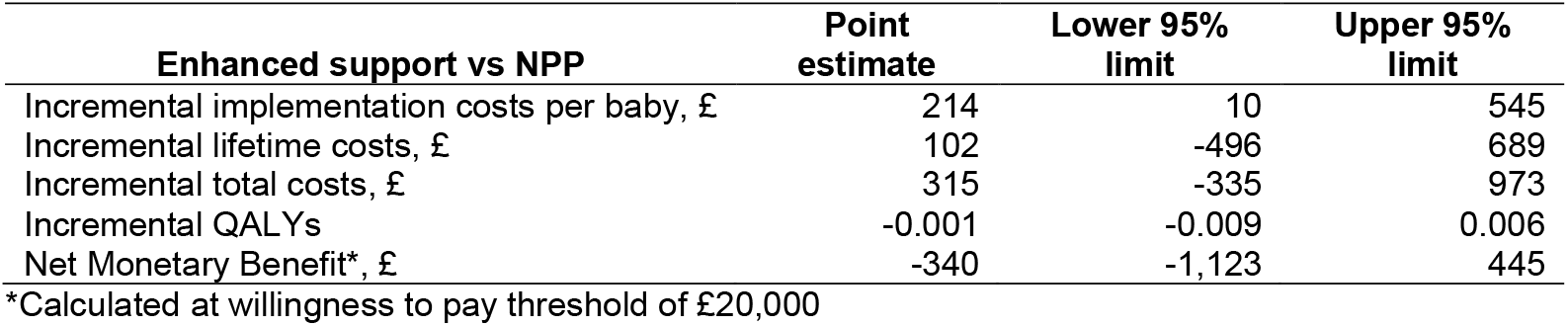
Probabilistic Analysis results of the enhanced support programme cost-effectiveness.

| **Enhanced support vs NPP** | **Point estimate** | **Lower 95% limit** | **Upper 95% limit** |
| --- | --- | --- | --- |
| Incremental implementation costs per baby, £ | 214 | 10 | 545 |
| Incremental lifetime costs, £ | 102 | -496 | 689 |
| Incremental total costs, £ | 315 | -335 | 973 |
| Incremental QALYs | -0.001 | -0.009 | 0.006 |
| Net Monetary Benefit*, £ | -340 | -1,123 | 445 |

\*Calculated at willingness to pay threshold of £20,000

Backfill funding for midwives and clinical champions allowed for on average 5.5 (enhanced support) and 1.7 (NPP) hours per week for PReCePT QI activities. The actual self-reported time spent per week over the first nine months was on target for the enhanced group (5.6 hours) but double the funded time for the NPP group (3.4 hours). This made the groups more similar than intended per protocol (Figure 2).

**Figure 2.**
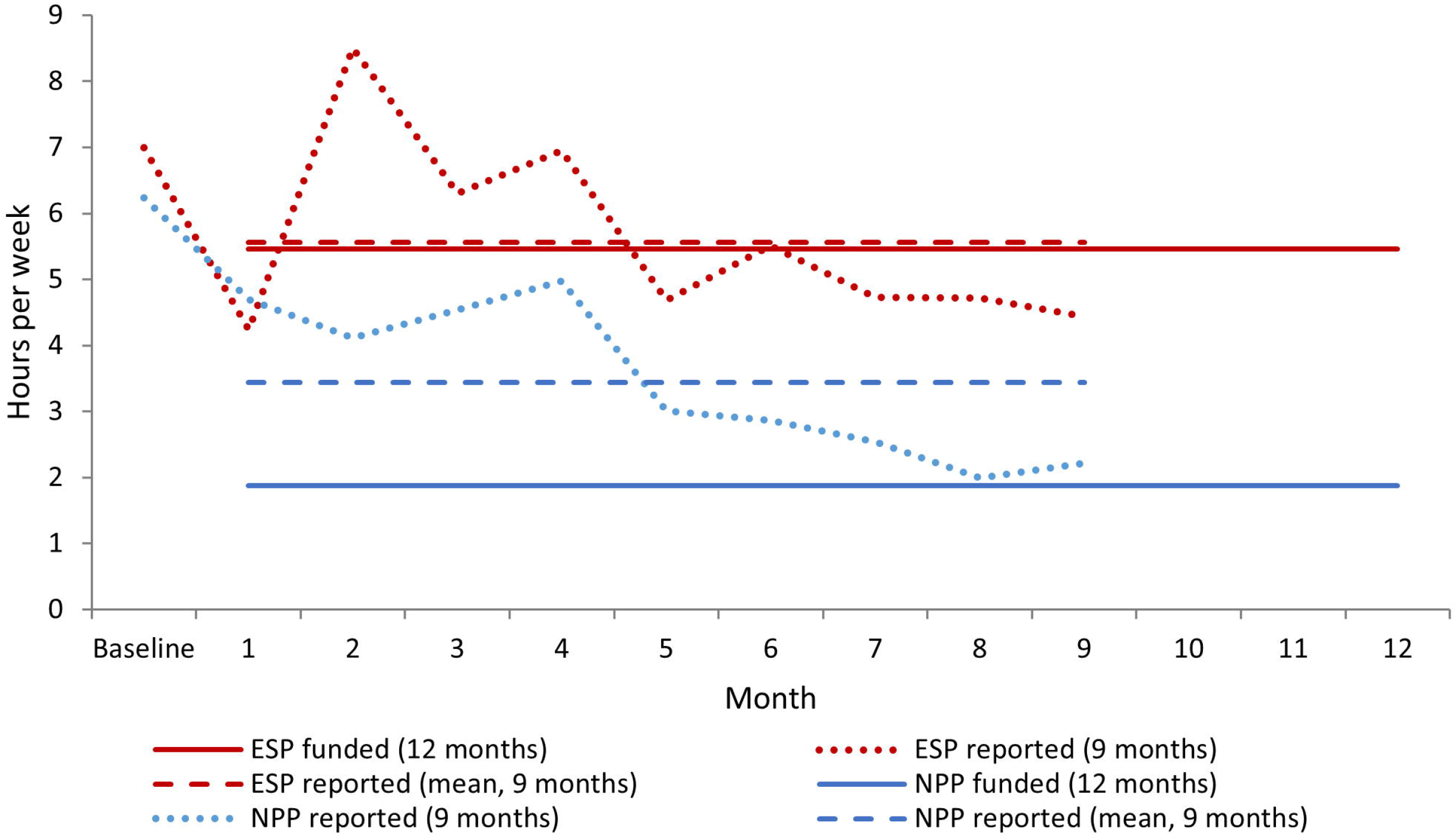

### Process Evaluation

Fifty-one participants were recruited from 29 units, representing ten out of the 12 AHSNs (Supplementary file 6). Twenty-two were lead midwives (two from the same unit), 14 lead obstetricians, and 15 lead neonatologists. Eighteen were from intervention units. Full results of the qualitative process evaluation are presented elsewhere.(21)

#### Similarities

Commitment to improving MgSO_4_ uptake was high among all units, encouraged by the NICE guidance, NNAP’s annual audit reporting, and the recent formation of the Maternity and Neonatal Safety Improvement Programme (MatNeo). Staff felt that the national character of the PReCePT programme and how the MgSO_4_ message was delivered was key to the intervention’s success.

The support structures in both groups were considered invaluable for understanding the project’s rationale, and for providing a ‘community of practice’ to share learning and ideas. For example: WhatsApp groups of midwives, AHSN and regional clinical leads; Twitter; and regional safety and improvement networks

Existing QI and implementation skills and capabilities varied across units, and so did training and support needs. Some NPP midwives had not received any QI training from AHSNs and would have liked more support. Some enhanced support midwives felt the intensive QI coaching had no added value over the support groups and activities they were already part of.

Both groups implemented core components of PReCePT QI. The “off the shelf” nature of the resources meant leads could pick and choose from ready-made tools according to what was appropriate to their setting. Implementation plans and resources were applied flexibly with local adaptations. Participants believed that improving collective knowledge and understanding through peer support and training, embedding MgSO_4_ in documentation, workflows, and clinical processes, and better data monitoring helped to embed MgSO_4_ use in clinical practice. The care of women in preterm labour and timely administration of MgSO_4_ required new routines that needed to be aligned with established responsibilities, and a vision for joint working across perinatal teams. PReCePT tried to make MgSO_4_ a shared goal and “everyone’s responsibility” by involving all members of the multi-professional care team in implementation activities.

#### Differences

Enhanced support learning events brought together midwifery, obstetric, and neonatology PReCePT leads, which helped develop relationships, and improved communication and collaboration. They collectively used their skills, abilities and networks to design and co-deliver PReCePT, with better support available to the lead midwife. Opportunities to come together helped counter the hectic, silo working typical in many clinical settings.

In contrast, the NPP model delivered support to champion midwives only. This encouraged the idea of PReCePT as a midwifery-specific, rather than a perinatal intervention. Team collaboration was more variable in NPP units and depended on the existing perinatal teamworking and safety culture. Several NPP midwives reported poor involvement and support from lead clinicians. NPP midwives with less contact with support structures or lower seniority/skillset, were less successful in delivering the full QI package and in overcoming challenges to implementation. NPP midwives were more likely to be left to manage the implementation alone, which often resulted in them having to work over their funded hours, and their time in practice often not being protected despite the backfill funding.

##### Impact of the COVID-19 pandemic

Implementers interviewed during the Coronavirus-19 pandemic reported several factors that could negatively impact on MgSO_4_ use and reporting: cessation of QI activities such as meetings and training, increased clinical pressures, staff shortages and reliance on untrained/agency staff.

## DISCUSSION

### Summary of results

MgSO_4_ uptake increased across maternity units in the NPP. Overall, the NPP was effective and cost-effective (full evaluation reported elsewhere(11)) The enhanced support model did not appear to improve MgSO_4_ uptake beyond the gains seen in the standard NPP support model. This indicates that the enhanced support would not be justified on a universal national scale. However, if part of the success of the NPP was due to the goodwill of staff delivering more than their funded time, these implementation costs should not be underestimated in future improvement programmes. Other similarities and networking connections between the groups reported in the process evaluation may have also contributed to the lack of observable difference.

Midwives’ success depended on staffing pressures, QI capacities, perinatal teamworking culture, and access to support (particularly the skills, seniority, and networks of obstetricians and neonatologists). Perinatal teamworking was an important enabler for improving MgSO_4_ uptake, and for embedding the practices that drive uptake. Enhanced support was more able to integrate and mobilise all members of the perinatal team. This is particularly important in light of the recent independent review of maternity services (2022 Ockenden report(22)), which found that a root cause of poor perinatal care is tribalism and deficient teamwork. If an intervention can even indirectly improve teamwork, this is likely to have far-reaching benefits across a broad range of perinatal outcomes. This is explored further in a separate forthcoming publication.

#### Impact of COVID-19

A slight decrease in MgSO_4_ uptake between March to June 2020 was observed, which might correspond to the first peak of Coronavirus-19 and the first UK lockdown. Women may have delayed presentation at hospital due to infection contact concerns, resulting in missed opportunities to give MgSO_4_. Clinical pressures observed in the process evaluation may have contributed to lower MgSO_4_ uptake, or less consistent reporting of administration. Further analysis of data beyond June 2020 would be valuable to identify uptake trends throughout the pandemic and longer-term sustainability of the PReCePT programme.

### Strengths and limitations

This is the first national-scale RCT of a QI intervention in perinatal medicine. It benefitted from use of robust, high-quality, routinely collected data, and a cluster design to minimise contamination between trial arms. Results represent 40/153 maternity units (26%) across England. Each perinatal team was able to tailor methods of implementing the toolkit to fit their local context, indicating that this sort of improvement programme can be highly successful while allowing flexibility, adaptability, and personalisation.(23)

There were variations in implementation between units. This variation is a key element of QI and a normal feature of real-world interventions but can hinder clear comparison between groups. NPP staff put in more than their funded hours for PReCePT activities, making them more similar than planned to enhanced support units. This could have contributed to the lack of observable difference between the groups.

The confidence intervals were consistent with the possibility of a small advantage (up to three percentage points) associated with enhanced support. The potential for even a small advantage should be considered, given the substantial lifetime benefits of avoiding CP. However, it is debatable whether an intervention that delivered this small advantage would be considered clinically important, given that NNAP’s latest annual report and a systematic review(24, 25) indicate that across many audit measures, a background annual improvement of a few percentage points would be expected anyway. Moreover, our probabilistic analysis indicated less than 30% probability that enhanced support was cost-effective.

This was a pragmatic trial embedded within a national scaling-up programme. As is the case for many policy evaluations, the ‘ideal’ direction of research evidence preceding and guiding policy decisions was not possible. The NPP was already being rolled-out and evaluation had to occur alongside it (or not at all). This can introduce challenges(26). Between trial design and trial start, the study landscape had changed significantly. The baseline rate of MgSO_4_ uptake had considerably increased, meaning that large differences between the groups, as observed in the pilot study, would not be achievable. This (as well as budget and eligibility criteria constraints on the sample size) limited the power of the study to detect small differences. However, as noted above, the confidence intervals around the null effect estimate are reasonably narrow, and we do not think it is likely they contain a difference that would be important for policy decisions on this subject. Given the cost difference between the two support models, evidence so far indicates that NPP-level support can be recommended over a more intensive support model. Longer-term analysis would, however, be valuable to identify any differences in sustainability of uptake. This is particularly indicated from the finding that perinatal teamwork tended to be better developed in enhanced support units, and better teamwork across staff groups is likely to be associated with greater sustainability of improvements, and benefits to other perinatal outcomes.

### Comparison with the literature

Uptake varies internationally, from 0%-12.3% in Europe(27) and 43.0% in Canada (2011-2015).(28) A clinician-led QI programme (single centre, Adelaide, Australia) reported increased uptake from 63% to 86% (2018-2021).(29) The programme included the establishment of a QI team, training and use of plan-do-study-act cycles. In Canada, MAG-CP was implemented (11 tertiary perinatal centres) and resulted in an absolute increase in uptake from 2.0% (2005-2010) to 46.3% (2011-2015).(30) This included educational rounds, focus group discussions and surveys of barriers and facilitators, on top of a national guideline and an online e-learning module.(30) Studies have also evidenced the feasibility of implementing MgSO_4_ clinical protocols in maternity units.(28, 29, 31) Their introduction in a tertiary hospital in the US resulted in a 73.9% absolute increase in uptake from 20% in 2007-2008 to 93.9% in 2011.(31) A smaller increase was seen in a French tertiary hospital, from 76% in 2011 to 87.5% in 2012.(28)

## Conclusion

The proportion of women given MgSO_4_ for preterm birth increased over the study period. The standard support model in the National PReCePT Programme overall was effective and cost-effective, and investing additional resources did not appear to benefit to MgSO_4_ uptake further. However, assessing individual hospitals’ specific support needs to tailor implementation may help to achieve greater uptake, and future quality improvement programs should not underestimate the staff time involved in the initial learning and implementation of better practice. The potential for indirect benefits such as better team working, and the positive impact this would have on services overall, should be explored further.

## Data Availability

Anonymised individual-level data for this study comes from the NNRD. Our data sharing agreement with the NNRD prohibits sharing data extracts outside of the University of Bristol research team.

## Other trial information

IRAS number 242419, ISRCTN 40938673, Trial Sponsor’s reference CH/2017/6417.

## Funding

The Health Foundation funded this trial (Funder’s reference 557668). The funders were not involved in study design, conduct, data collection, analysis, interpretation, or writing of this manuscript. This research was supported by the National Institute for Health and Care Research (NIHR) Applied Research Collaboration West (NIHR ARC West, core NIHR infrastructure funded: NIHR200181). The views expressed in this article are those of the author(s) and not necessarily those of the NIHR or the Department of Health and Social Care.

## Contributors

KL, BO, and JD conceptualized the trial; KL and BO led the funding application to the Health Foundation supported by JD; KL is Chief Investigator and BO is Co-Chief Investigator. TP, MTR, SR, WH, and HM led the design and analysis plan. ET advised on methodology. PC and EMH were trial managers. HE, MTR, RM, CSR and PC acquired NNRD and questionnaire data. HE, MTR and RM conducted the effectiveness analysis. CSR, HM and WH conducted the cost-effectiveness analysis. CPM, TS and SR conducted qualitative data collection and analysis. HE, MTR, CSR, CPM and RM wrote the original manuscript and contributed equally to the paper. All authors reviewed and edited the manuscript and approved the submission. KL is guarantor.

## Ethics and regulatory considerations

The UK National Health Service Health Research Authority (NHS HRA) approved the conduct of the trial (HRA ID 242419) and gave authorisation that it did not require Research Ethics Committee approval as a low-risk study involving NHS staff who had given consent as participants and used pseudonymised patient data.

## Conflicts of interest

All authors in this manuscript have no conflict of interest to declare aside from funding from NIHR ARC West, AHSN, NHS England and The Health Foundation as detailed above. We declare that the study management group have no competing financial, professional, or personal interests that might have influenced the study design or conduct.

## Acknowledgements

Public and Patient Involvement for this trial built on the involvement work in the PReCePT pilot study.(10) This used a co-design and co-production approach including a partnership with BLISS, a support organisation for mothers experiencing preterm births, and two mothers who had experienced preterm births, Elly Salisbury and Monica Bridge who were involved in trial design and delivery (at the learning events) and were part of the Trial Steering Committee.

We also acknowledge The Health Foundation and the West of England Academic Health Science Network (in particular Natasha Swinscoe and Ellie Wetz) for their support and guidance, the AHSN Network (in particular Gary Ford for leadership and guidance), Anna Burhouse for her continued input and inspiration, QI Coaches Noshin Menzies, Vardeep Deogan and Hannah Bailey, Jo Bangoura for producing the PReCePT QI toolkit, and all local champions who were instrumental in applying the QI training from learning events to their local perinatal teams.

## Data sharing

Pseudonymised individual-level data for this study comes from the NNRD. Our data sharing agreement with the NNRD prohibits sharing data extracts outside of the University of Bristol research team. Copies of the NNRD data dictionary and the full study protocol are available online(12) and copies of the Statistical Analysis Plan are available on the University of Bristol Research Information System (https://research-information.bris.ac.uk/en/projects/precept-study-a-cluster-randomised-trial-evaluating-the-impact-of).

## Notes

### Competing Interest Statement

The authors have declared no competing interest.

### Clinical Trial

ISRCTN 40938673

### Clinical Protocols

https://pubmed.ncbi.nlm.nih.gov/34031151/

### Funding Statement

The Health Foundation funded this trial (Funder reference 557668). The funders were not involved in study design, conduct, data collection, analysis, interpretation, or writing of this manuscript.
This research was supported by the National Institute for Health and Care Research (NIHR) Applied Research Collaboration West (NIHR ARC West, core NIHR infrastructure funded: NIHR200181). The views expressed in this article are those of the author(s) and not necessarily those of the NIHR or the Department of Health and Social Care.

### Author Declarations

The UK National Health Service Health Research Authority (NHS HRA) approved the conduct of the trial (HRA ID 242419) and gave authorisation that it did not require Research Ethics Committee approval as a low-risk study involving NHS staff who had given consent as participants and used anonymised patient data.

### Summary of Updates

Minor revisions to description of qualitative findings and discussion section.

