## Supplementary for "Evaluation of standard and enhanced quality improvement methods to increase the uptake of magnesium sulfate in preterm deliveries for the prevention of neurodisability (PReCePT Study): a cluster randomized controlled trial"

### Supplementary File

#### 1. Description of trial groups

| | NPP support (Control) | Enhanced support (Intervention) |
|---|---|---|
| PReCePT QI toolkit | Clinical guidance;<br>Pre-term labour proforma template;<br>Staff training presentations;<br>Parent leaflet;<br>Posters for display on the unit to raise staff awareness;<br>A QI Learning Log;<br>Project Dashboard;<br>Pens, magnets, lanyards and other aide-mémoires to promote $MgSO_4$ to unit staff (if purchased) | As per standard support group |
| QI training | Local level (AHSN or unit level) QI training and guidance to adapt materials for local use, cascaded from AHSN | As per standard support group |
| Regional support | Support from a AHSN level clinical lead (obstetrician and neonatologist) and AHSN lead | As per standard support group |
| Local clinical champion | Local obstetrician and neonatologist identified by unit to guide and oversee local implementation | As per standard support group (named as joint PI, at discretion of local site) |
| Funded time for local midwife champion | Funded time of up to 90 hours per unit (on average 2 hours per week) | As per standard support, PLUS funding for up to 90 extra hours backfill, on average over 12 months, to enable the midwife to embed the QI toolkit within their team |
| Funded time for local neonatologist champion | None | Funded time for a local neonatologist PI , working on average 0.5 PA (2 hours) per week over 12 months, to provide clinical leadership in local unit (fixed term contract or secondment from an NHS organisation)<br>0.5 PA backfill may be split with obstetrician PI, at discretion of local site |
| QI coaching | None | Structured coaching in local unit from an experienced QI coach. To include:<br>First visit where the QI coach will work with local unit to create a bespoke implementation plan;<br>Telephone coaching in liaison with the local champion(s), with occasional face to face visits as logistics permit;<br>Ongoing dedicated support to help embed the QI toolkit within local unit;<br>Final visit to support local unit to tie up data collection and plan for ongoing sustainability |
| Learning events | Shared learning events between AHSN managers leading the NPP in their area | Funding for up to three members of staff from local unit to attend three learning events. These bespoke learning events will be held every 2-3 months during the period of implementation and will bring together teams from other Group 2 units to share their activity and learning on how they are implementing the PReCePT QI toolkit and working to address issues and challenges |
| Celebration event | None | Provision of an android tablet to be used by the local midwife champion to micro-coach colleagues, plus a small fund for purchasing study collateral (pens, magnets, lanyards, aide-mémoires), if required |
| Collateral funding | None | Funding for up to three members of staff from local unit to attend a celebration event which will bring together teams from all Group 2 units to wrap up the study and to share experiences, learning and success. |

#### 2. Study randomisation

Of the 80 eligible units, 48 consented to participate in the study. Of units eligible in wave one, 29 consented and eight were allocated to the intervention group and 16 to the control group. Five units were allocated as reserve, but these only represented two strata. Four control units withdrew due to change in unit readiness for NPP implementation and two of these units were replaced by reserve units. There were no reserve units available for the other two units that withdrew.

Of units eligible in wave two, 19 consented to participate in the study. There were, however, not enough consenting units to keep to the 2:1 control:intervention ratio for all strata. To keep the ratio for stratum 1, we randomised 1 unit to the intervention arm and 2 units to the control arm. Units in strata 2 and 3 were randomised to intervention and control arms using the 1:2 ratio as closely as possible, allocated independently for each stratum. We had 1 control and 1 intervention unit in strata 2 and 1 control and no intervention unit in strata 3. We oversampled for the last stratum, making use of all units available and had 4 intervention units and 8 control units. Six units were allocated to the intervention group and 13 to the control group. One intervention unit withdrew due to change in NPP implementation readiness.

**Table S1. Distribution of study units by Baseline $MgSO_4$.**

| Uptake | Enhanced support (intervention) | NPP standard support (control) | Units per strata |
|---|---|---|---|
| Stratum 1: 0-39% | 3 | 5 | 8 |
| Stratum 2: 40-49% | 2 | 5 | 7 |
| Stratum 3: 50-59% | 2 | 5 | 7 |
| Stratum 4: 60-≤70% | 6 | 12 | 18 |
| Total | 13 | 27 | 40 |

#### 3. Economic evaluation

**Table S2. Estimated lifetime costs and QALYs per patient associated with $MgSO_4$ treatment.**

| Type of birth | Perspective | Method | Cost, £[*] | Δcost, £* | QALYs | ΔQALYs |
|---|---|---|---|---|---|---|
| Imminent | Societal | $MgSO_4$ | 61,971 | -23,690 | 26.6 | 0.3 |
| | | No $MgSO_4$ | 85,661 | | 26.3 | |
| Threatened | Societal | $MgSO_4$ | 44,068 | -15,964 | 26.7 | 0.2 |
| | | No $MgSO_4$ | 60,032 | | 26.5 | |

[†] Based on Bickford et al (7)
[*]Cost estimates were converted to Pounds Sterling and inflated to 2019 prices

**Table S3. Point estimates, probability distributions, and source of parameter estimates used in the probabilistic analysis.**

| Parameter | Statistics | Estimate | Probability distribution | Source |
|---|---|---|---|---|
| Type of birth (Imminent) | n (%) | 1017/2493 (39%) | Beta distribution | Trial data |
| Probability $MgSO_4$ treatment (yes) – Control arm | | | | |
| $MgSO_4$ yes - Imminent | n (%) | 591/703 (84%) | Beta distribution | Trial data |
| $MgSO_4$ yes - Threaten | n (%) | 805/989 (81%) | Beta distribution | Trial data |
| Incremental effectiveness ESP vs NPP | | | | |
| $MgSO_4$ yes - Imminent | OR (se) | 0.99 (0.03) | LogNormal | Logistic regression |
| $MgSO_4$ yes - Threaten | OR (se) | 0.99 (0.03) | LogNormal | Logistic regression |
| Incremental lifetime QALYs | | | | |
| $MgSO_4$ yes - Imminent | Mean (se*) | 0.3 (0.06) | Beta distribution | Bickford et al[1] |
| $MgSO_4$ yes - Threaten | Mean (se*) | 0.2 (0.04) | Beta distribution | Bickford et al[1] |
| Incremental lifetime costs | | | | |
| $MgSO_4$ yes - Imminent | Mean (se*) | £-23,690 (-4,738) | Gamma distribution | Bickford et al[1] |
| $MgSO_4$ yes - Threaten | Mean (se*) | £-15,964 (-3,193) | Gamma distribution | Bickford et al[1] |
| NPP Implementation costs | | | | |
| $MgSO_4$ yes - Imminent | Mean (sd) | £94 (65) | Gamma distribution | Trial data |
| $MgSO_4$ yes - Threaten | Mean (sd) | £97 (67) | Gamma distribution | Trial data |
| $MgSO_4$ no - Imminent | Mean (sd) | £97 (65) | Gamma distribution | Trial data |
| $MgSO_4$ no - Threaten | Mean (sd) | £102 (74) | Gamma distribution | Trial data |
| ESP additional implementation costs[#] | | | | |
| $MgSO_4$ yes - Imminent | Mean (sd) | £262 (267) | Gamma distribution | Trial data |
| $MgSO_4$ yes - Threaten | Mean (sd) | £285 (298) | Gamma distribution | Trial data |
| $MgSO_4$ no - Imminent | Mean (sd) | £233 (205) | Gamma distribution | Trial data |
| $MgSO_4$ no - Threaten | Mean (sd) | £318 (287) | Gamma distribution | Trial data |

*Standard Errors were assumed to be 20% of their point estimates

[#]The total cost of the ESP model (intervention arm) is the result of the sum of NPP and the ESP additional implementation costs

NPP National PReCePT Programme

ESP Enhanced Support Programme

**Table S4. Mean unit-level funded costs for the enhanced support programme and NPP.**

| Category | Intervention (n=13) | Control (n=27) | difference |
|---|---|---|---|
| | ESP | NPP | |
| Central, £ | 780 | 780 | 0 |
| AHSN, £ | 2,492 | 2,764 | -272 |
| Clinical time, £ | 10,000 | 2,500 | 7,500 |
| QI coaches, £ | 3,254 | 0 | 3,254 |
| Other[†], £ | 6,387 | 0 | 6,387 |
| Total | 22,913 | 6,044 | 16,869 |

[†] Including project management, learning events, and miscellaneous

**Table S5. Mean lifetime QALYs and costs per baby by type of birth and trial arm.**

| | ESP (Intervention) | | | | | NPP (Control) | | | | |
|---|---|---|---|---|---|---|---|---|---|---|
| | Imminent | | Threaten | | | Imminent | | Threaten | | |
| | $MgSO_4$ Yes | $MgSO_4$ No | $MgSO_4$ Yes | $MgSO_4$ No | Total | $MgSO_4$ Yes | $MgSO_4$ No | $MgSO_4$ Yes | $MgSO_4$ No | Total |
| Lifetime costs | 61,971 | 85,661 | 44,068 | 60,032 | 54,554 | 61,971 | 85,661 | 44,068 | 60,032 | 54,416 |
| Mean Implementation costs | 350 | 315 | 381 | 425 | 372 | 94 | 97 | 97 | 102 | 97 |
| QALYs | 26.60 | 26.30 | 26.70 | 26.50 | 26.62 | 26.60 | 26.30 | 26.70 | 26.50 | 26.62 |

NPP National PReCePT Programme
ESP Enhanced Support Programme

**Table S6. Probabilistic Analysis results of the enhanced support programme cost-effectiveness.**

| ESP vs NPP | Deterministic analysis |
|---|---|
| Incremental implementation costs per baby, £ | 276 |
| Incremental lifetime costs, £ | 137 |
| Incremental total costs, £ | 413 |
| Incremental QALYS | -0.002 |
| Net Monetary Benefit*, £ | -447 |

*****used a willingness-to-pay threshold of £20,000 per QALY
ESP Enhanced Support Programme
NPP National PreCePT Programme

**Figure S1. Cost-effectiveness plane of PReCePT Enhanced Support versus National PreCePT Programme Support.**

The graph displays results of Monte Carlo simulations with 10 000 iterations using the value ranges and distributions presented in Table S3. The horizontal axis represents the difference in effect measures in Quality Adjusted Life Years (QALYs) between the enhanced support versus the standard support; and the vertical axis represents the difference in cost. Datapoints falling top left quadrant indicate that the enhanced support is less effective and more costly than the standard support. Datapoints falling the top right quadrant indicate that the enhanced support is more effective and more costly than the standard support. Datapoints falling bottom right quadrant indicate that the enhanced support is more effective and less costly than the standard support.

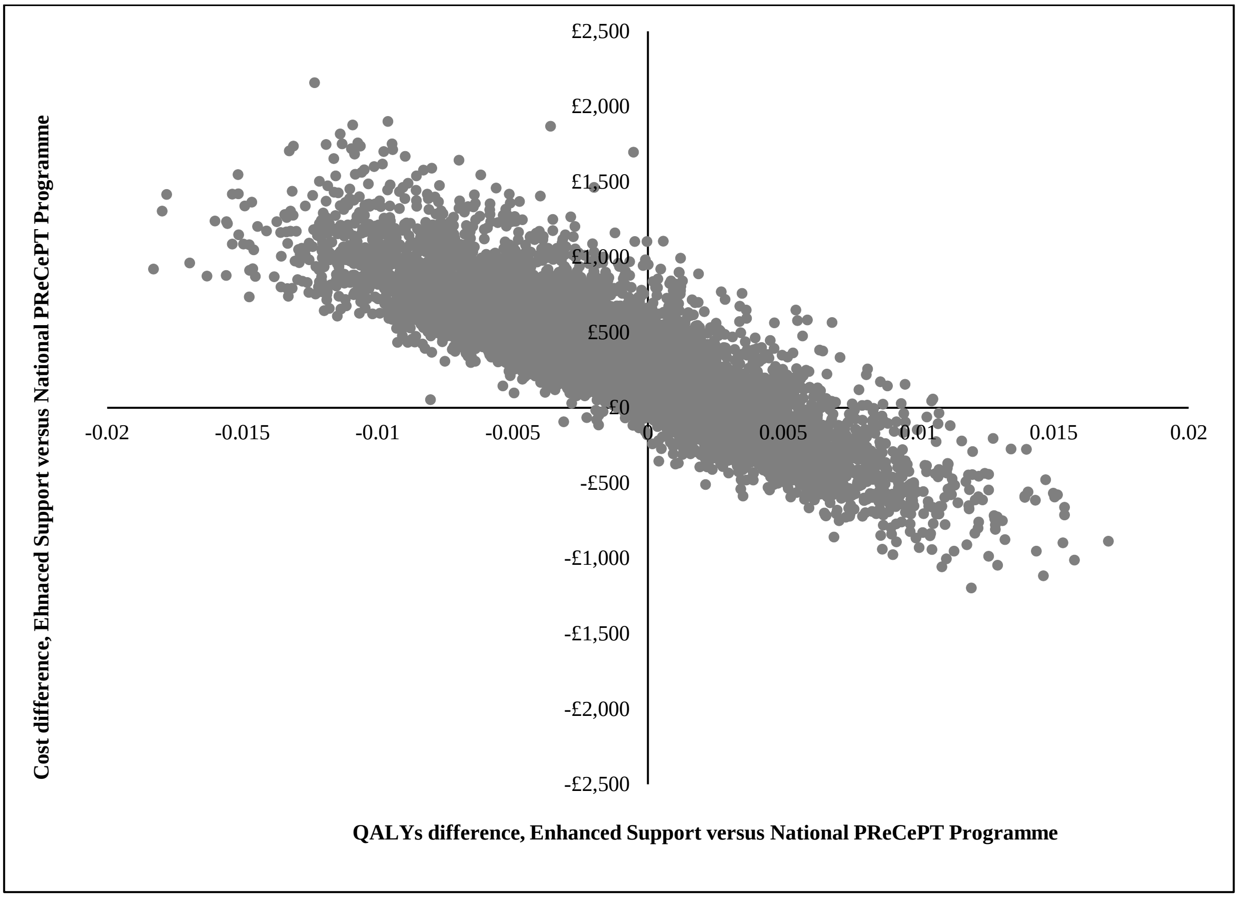

**Figure S2. Cost-effectiveness acceptability curve of PReCePT Enhanced Support versus National PReCePT Programme Support.**

The curve shows the probability of enhanced support being cost-effective at different cost-effectiveness threshold values.

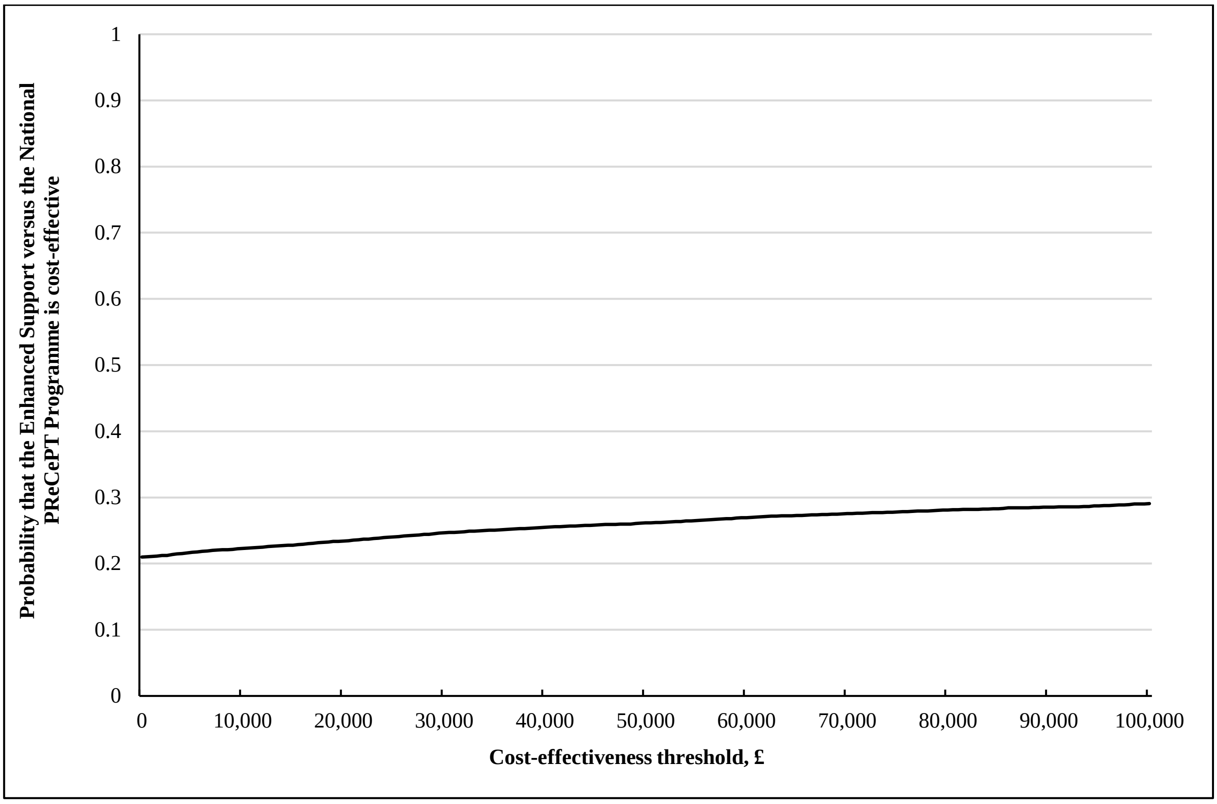

#### 4. MgSO4 over time

**Figure S3: MgSO4 uptake and data completeness over time**

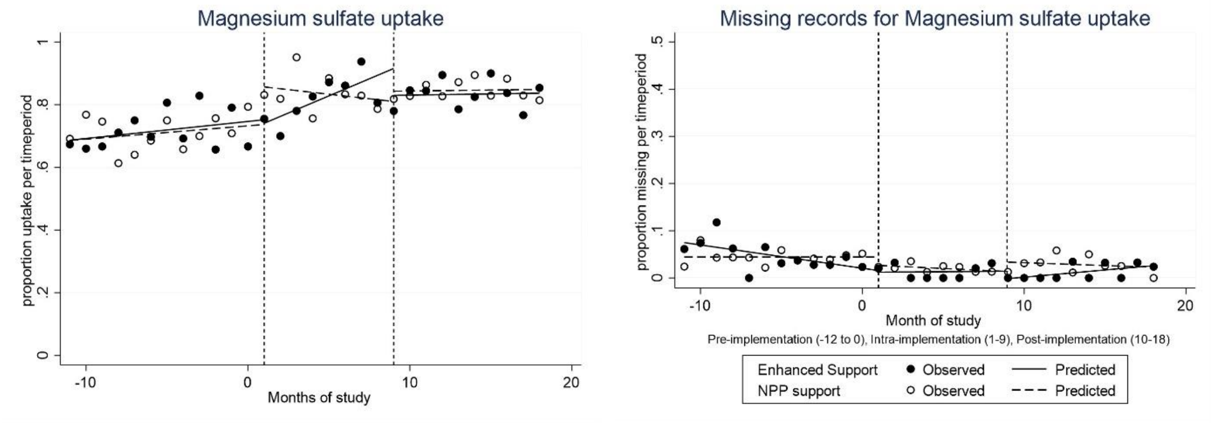

#### 5. Qualitative interviews topic guide

**PReCePT Study Staff Interview Topic Guide**

Participant groups (unit lead midwife, unit lead neonatologist, unit lead obstetrician, doctor in training, staff midwife)

***1. Background***

- Provide an overview of interview purpose, intended use for the interview data, measures to protect confidentiality and anonymity, permission to audio record interview and take notes,
- Any other questions before we start? (start recording)
- Receive verbal consent if this has not already been done

***2. Introductions***

- Participants' job title, time in post, role and responsibilities in the unit
- Role in PReCePT Study

***3. Opening***

- What has been your experience of PReCePT at your unit? Can you briefly talk me through the process you follow to administer $MgSO_4$? In your view, how has it been going?

***4. Specific questions about the PReCePT QI intervention***

(these will vary for each group of participants depending on their role in the study and whether the participant works in a unit which is 'enhanced support' (intervention arm) or 'standard support' (control arm))

- Overall, what are your views of the PReCePT QI Toolkit and implementation guide?
- Could I take you through the PReCePT QI Toolkit and ask you to comment on the usefulness of each?
    - Introduction to PReCePT Evidence (Essential resources)
    - Summary of Key Research (Essential resources)
    - Clinical Guidance for the Management of Suspected Preterm Labour (Essential resources)
    - PReCePT Magnesium Sulphate Quick Reference (Essential resources)
    - PReCePT Infographic (Essential resources)
    - Parent Information Leaflet (Essential resources)
    - Staff "PReCePT Training" Presentation (Essential resources)
    - Midwife Lead Role Description (Essential resources)
    - Regional Neonatal Lead Role Description (Essential resources)
    - Maternity Unit Obstetrician Lead Role Description (Essential resources)
    - Poster: Think Magnesium Sulphate Too (Strongly Recommended resources)
    - The PReCePT Dashboard and how to use it (Strongly Recommended resources)
    - PReCePT Management of Preterm Labour Proforma (Optional resources)
    - PReCePT Magnet Instructions (Optional resources)
    - QI Learning Log (Optional resources)
    - Other you've used in your unit e.g. pens, lanyards?
- Did you amend any of the materials to suit your local needs?
- Would you have preferred something different?
- Could I take you back to the launch event and the QI training you received?
    - How useful did you find it? For your role in PReCePT / your role overall e.g. applied in other areas of work?
    - What did you like about it/ didn't like about it? Would you like anything to be different?
- What coaching did you receive?
    - How frequently have you been in touch with the QI coaches? Via phone, emails, social media, face to face?
    - Was this contact instigated by the coaches or your unit? What were the reasons for (non)engagement?
    - How useful did you find it?
    - What did you like about it/didn't like about it? Would you like anything to be different?
    - Do you draw support from any outside resources? E.g. social media groups, twitter, other? Which ones?

***5. Specific questions about the PReCePT QI implementation***

(these will vary for each group of participants depending on their role in the study and whether the participant works in a unit which is 'enhanced support' (intervention arm) or 'standard support' (control arm))

- In your view, to what extent did your unit adhere or not to the PReCePT QI training [when administering $MgSO_4$]? What were the reasons? [Have you adapted it in any way to fit your setting?]

- Were there any factors that may have influenced the implementation and observed outcomes; such as organisational changes, staff shortages? Were there any professional, organisational or cultural issues that may have affected implementation? How has the collaboration between obstetrics and postnatal staff been?

- To what extent were staff engaged in PReCePT? Were there some staff more engaged than others? Were there opportunities for involvement? For making suggestions, comments and changes?

- Were patients involved in the implementation of PReCePT? Has a local PPI representative been involved? To what extent and in what way?

- To what extent did PReCePT as a model for quality improvement fit in or interfered with the daily practice?  Has it become embedded in usual practice or has it been experienced as disruptive? Why?

- To what extent did the unit/hospital/trust leadership support PReCePT 2?

***6. Closing***

- Thank you for your feedback. Is there anything else you would like to tell me for the evaluation?
- Provide information about when the findings from the study will be made available, publications plans, current issues and developments.

#### 6. A description of units participating in qualitative interviews

| Unit Code | AHSN Code | Arm | 2017 $MgSO_4$ uptake | 2019/20 $MgSO_4$ uptake | Type of unit (No of eligible mothers in 2017) | CQC patient safety rating | CQC overall rating | Number of interviews conducted |
|---|---|---|---|---|---|---|---|---|
| 12 | **A** | Standard support | 34.1% | 70.83% | Non-tertiary unity (N=41) | Requires improvement | Good | 1 |
| 5 | **B** | Enhanced support | 65.0% | 100% | Non-tertiary (N=40) | Good | Good | 2 |
| 15 | | Standard support | 68.0% | 78.8% | Tertiary (N=75) | Requires improvement | Good | 3 |
| 16 | | Standard support | 50.0% | 89.0% | Non-tertiary (N=10) | Good | Good | 1 |
| 1 | **C** | Standard support | 41.2% | 100% | Non-tertiary (N=17) | Good | Good | 2 |
| 9 | | Standard support | 61.9% | 82.2% | Non-tertiary (N=21) | Requires Improvement | Good | 1 |
| 14 | | Standard support | 40.0% | 67.0% | Non-tertiary (N=15) | Good | Good | 3 |
| 23 | **D** | Enhanced support | 27.3% | 100% | Non-tertiary (N=11) | Requires improvement | Good | 2 |
| 24 | | Standard support | 39.7% | 76.6% | Tertiary (N=58) | Requires improvement | Good | 3 |
| 28 | | Standard support | 57.1% | 38.9% | Non-tertiary (N=14) | Good | Good | 1 |
| 29 | | Enhanced support | 67.0% | 87.0% | Tertiary (N=81) | Good | Good | 1 |
| 13 | **E** | Standard support | 67.5% | 90.3% | Tertiary (N=80) | Good | Good | 3 |
| 33 | | Standard support | 60.0% | 100% | Non-tertiary (N=20) | Good | Good | 2 |
| 32 | **F** | Enhanced support | 70.0% | 100% | Non-tertiary (N=10) | Good | Good | 1 |
| 34 | | Standard support | 64.3% | 75.0% | Non-tertiary (N=14) | Good | Good | 1 |
| 4 | **G** | Enhanced support | 41.2% | 100% | Tertiary (N=53) | Good | Good | 2 |
| 22 | | Standard support | 62.2% | 74.5% | Non-tertiary (N=45) | Good | Requires Improvement | 1 |
| 6 | **H** | Enhanced support | 42.1% | 76.5% | Tertiary (N=57) | Good | Outstanding | 1 |
| 7 | | Enhanced support | 60.0% | 55.6% | Non-tertiary (N=15) | Good | Good | 3 |
| 8 | **K** | Enhanced support | 55.6% | 75.6% | Non-tertiary (N=18) | Requires improvement | Good | 1 |
| 11 | | Standard support | 38.5% | 75.6% | Non-tertiary (N=13) | Good | Good | 2 |
| 17 | | Standard support | 47.5% | 85.2% | Non-tertiary (N=40) | Good | Good | 2 |
| 19 | | Standard support | 44.4% | 79.4% | Non-tertiary (N=27) | Requires improvement | Good | 1 |
| 20 | | Standard support | 7.1% | 83.3% | Non-tertiary (N=14) | Good | Good | 2 |

| | | | | | | | | |
|---|---|---|---|---|---|---|---|---|
| 36 | **L** | Standard support | 65.0% | 100% | Non-tertiary (N=20) | Requires improvement | Good | 2 |
| 37 | | Standard support | 70.9% | 91.3% | Tertiary (N=55) | Requires improvement | Good | 1 |
| 31 | **M** | Enhanced support | 68.9% | 85.7% | Tertiary (N=74) | Good | Outstanding | 3 |
| 38 | | Standard support | 61.5% | 60.0% | Non-tertiary (N=13) | Good | Good | 1 |
| 40 | | Standard support | 68.4% | 87.0% | Non-tertiary (N=19) | Requires improvement | Good | 2 |

1. Bickford CD, Magee LA, Mitton C, et al. Magnesium sulphate for fetal neuroprotection: a cost-effectiveness analysis. *BMC Health Services Research* 2013; **13**(1): 527.
